# Online training in manuscript peer review: a systematic review

**DOI:** 10.1101/2022.09.02.22279345

**Authors:** Jessie V. Willis, Kelly D. Cobey, Janina Ramos, Ryan Chow, Jeremy Y. Ng, Mohsen Alayche, David Moher

## Abstract

**Background:** Peer review plays an integral role in scientific publishing. Despite this, there is no training standard for peer reviewers and review guidelines tend to vary between journals. The purpose of this study was to conduct a systematic review of all openly available online training in scholarly peer review and to analyze their characteristics.

**Methods:** MEDLINE, PsycINFO, Embase, ERIC, and Web of Science were systematically searched. Additional grey literature searches were conducted on Google, YouTube, university library websites, publisher websites and the websites of peer review related events and groups. All English or French training material in scholarly peer review of biomedical manuscripts openly accessible online on the search date (September 12, 2021) were included. Sources created prior to 2012 were excluded. Screening was conducted in duplicate in two separate phases: title and abstract followed by full text. Data extraction was conducted by one reviewer and verified by a second. Conflicts were resolved by third-party at both stages. Characteristics were reported using frequencies and percentages. A direct content analysis was preformed using pre-defined topics of interest based on existing checklists for peer reviewers. A risk of bias tool was purpose-built for this study to evaluate the included training material as evidence-based. The tool was used in duplicate with conflicts resolved through discussion between the two reviewers.

**Results:** After screening 1244 records, there were 43 sources that met the inclusion criteria; however, 23 of 45 (51%) were not able to be fully accessed for data extraction. The most common barriers to access were membership requirements (n = 11 of 23, 48%), availability for a limited time (n = 8, 35%), and paywalls with an average cost of $99 USD (n = 7, 30%). The remaining 20 sources were included in the data analysis. All sources were published in English. Half of the sources were created in the last five years (n = 10, 50%). The most common training format was an online module (n = 12, 60%) with an estimated completion time of less than one hour (n = 13, 65%). The most frequently covered topics included how to write a peer review report (n = 18, 90%), critical appraisal of data and results (n = 16, 80%), and a definition of peer review (n = 16, 80%). Critical appraisal of reporting guidelines (n = 9, 45%), clinical trials (n = 3, 15%), and statistical analysis (n = 3, 15%) were less commonly covered. Using our ad-hoc risk of bias tool, four sources (20%) met our criteria for evidence-based.

**Conclusion:** Our comprehensive search of the literature identified 20 openly accessible online training materials in manuscript peer review. For such a crucial step in the dissemination of literature, a lack of training could potentially explain disparities in the quality of scholarly publishing. Future efforts should be focused on creating a more unified openly accessible online manuscript peer review training program.

## INTRODUCTION

Peer review is often considered to be of vital importance to the credibility of scholarly publishing^1-3^. It encompasses the standard practice in which handpicked experts, or “peers”, critically appraise a submitted manuscript for any concerns that would make it unsuitable for publication^4^. Peer reviewers are expected to provide constructive feedback to the authors and, if asked, a recommendation to the journal editor on whether to reject or accept the paper^5^.

Despite being the accepted gold standard, peer review is not free from criticism and skepticism^6,7^ with inquiries into improving and innovating peer review at the forefront of discussion^8,9^. Previous studies have shown peer review to lack transparency, reproducibility, and consistency^10-14^. Since peer review is typically conducted internally by each journal, there is no standard training, objective guidelines, or formal evaluation done for reviewers^15,16^. A recent study demonstrated that journal-provided reviewer guidelines varied greatly between journals^17^.

The current state of peer review requires a feasible intervention that can improve quality standards and provide uniformity^18^. Specifically, an established training standard for peer reviewers could address many of the current limitations^19^. A standard training process could provide assurance that all peer reviewers are knowledgeable on topics core to functioning as a reviewer, including but not limited to selective reporting bias and “spin”^20,21^. Unfortunately, data currently available on training interventions is small-scale and shows insignificant benefit^22-25^. Additionally, these studies were published before the popularization of online training, which would be a much more convenient format for journals or institutions to implement. Additionally, recent surveys have shown training to be highly desired, especially by early career researchers (ECRs)^26,27^. A 2018 online survey conducted by Publons found that 88% of respondents believe training is important for ensuring credibility of peer review^28^.

Given the above, we performed a systematic review of all openly available online training material for peer review of biomedical manuscripts. An analysis of the characteristics of the existing corpus of training material could potentially assist in the development of future standardized training programs.

## METHODS

This article was written in accordance with the PRISMA guidelines for systematic reviews^29^. The study protocol was registered on Open Science Framework prior to data collection (https://osf.io/x5yc9) and was used as a direct reference for the creation of this manuscript^30^.

### 1. Data sources and searches

The search strategy was developed in collaboration between two members of the review team (JVW, JYN) and a medical research librarian (LS). Another medical librarian (RS) peer reviewed the search strategy using the Peer Review of Electronic Search Strategies (PRESS) Checklist^31^. Our search of databases was not restricted by language, as we wished to record the existence of training in other languages. The following databases were searched through the OVID platform: MEDLINE, PsycINFO, Embase and ERIC on September 12, 2021. In addition, Web of Science was searched on September 13, 2021. The OVID search strategy is reported in Appendix 1. A forward and backward citation analysis was conducted of sources included at the data extraction stage. Preprint servers (OSF and MedRxiv) and PROSPERO were also searched to identify similar systematic reviews. Grey literature searches were conducted of Google, YouTube, university library websites, publisher websites, and websites of peer review related groups/events. The full grey literature search strategy can be found in Appendix 2.

### 2. Eligibility criteria

Included sources were online educational material intended for biomedical manuscript peer reviewers available and openly accessible on the search date, published in either English or French, and created after January 1, 2012. The rationale for the cut-off year was based on the year the Publons website (https://publons.com/, Clarivate Analytics) was launched. The launch of Publons marked a significant innovation in peer review and a call for higher quality reviewing^32^. Additionally, Publons had developed the earliest freely accessible online training material for peer review recognized by the authors. For this reason, it was thought that any high-quality and relevant training in the current state of peer review would have been created after 2012. Exclusion criteria included sources describing other types of peer review beyond that at academic journals (e.g., grant review, review for tenure and promotion) or publications on peer review that are opinion-based (e.g., commentary, opinion articles). We recorded the existence of sources that were inaccessible (e.g., paywall, membership) or in a language other than English or French, but these were excluded from data analysis.

### 3. Screening process

Search results were imported into DistillerSR (Evidence Partners Inc., Ottawa, Canada), which was used as the software for removing duplicates, screening, and data extraction. Screening was done in an expedited process whereby all articles were screened by one reviewer and only excluded articles were verified by a separate independent reviewer. The screening was conducted in two stages: title and abstract followed by full text. Conflicts were resolved by consensus or through a third-party screener at both stages. Screening questions can be found in Appendix 3.

### 4. Data extraction and internal validity assessment

Data extraction was performed in DistillerSR. The data extraction form was pilot tested by each of the two reviewers with 10 records each. Data extraction was completed in duplicate by two reviewers independently. Conflicts were resolved by consensus discussion between the two reviewers or by a third-party. The data extraction items can be found in Appendix 4.

In traditional systematic reviews (i.e., assessing the effects on an intervention) risk of bias assessment is considered a best practice. There is no risk of bias tools for methodological reviews. We developed our own internal validity tool following previous efforts^34^. The questions were pilot tested by our data extractors. There were six items on our tool. Briefly, these were 1) having representation of greater than one stakeholder group in the author list, 2) reporting data gathering (i.e., conducting a survey) for development of the training, 3) reporting pilot testing, 4) having clear learning objectives, 5) having a method of self-testing or evaluation, and 6) having a method of providing feedback. For each selected source, this questionnaire was completed by two independent reviewers in Excel, and conflicts were resolved by consensus (see Appendix 5 for full internal validity tool). We considered a training ‘evidence-based’ if four (of the six) questions were answered affirmatively.

### 5. Data synthesis and analysis

The training sources were evaluated qualitatively and compared against each other for overall content. Results were reported in table format and with frequencies and percentages listed. A directed content analysis was done with predefined codes based on existing checklists for peer reviewers, reporting guidelines and expert opinion (DM)^33^.

## RESULTS

### 1. Deviations from our protocol

We had initially defined “course registration” as an exclusion criterion as it could potentially be a barrier to access. Additionally, we had believed it would be infeasible to register for each training material in the data extraction phase. However, a registration requirement was common with a large portion of references blocked by registration alone. It was therefore decided to include these sources. Two independent reviewers registered for these sources to perform full data extraction.

### 2. Study selection

We identified 1058 sources with the database search and 487 sources with the grey literature search. Following duplicate removal, a total of 1244 records were screened, and 43 sources were identified as eligible. Of these eligible sources, there were 23 (51%) that had limited access and could not be included in data extraction. One of these sources had an advanced version blocked by paywall but a free basic version that was added to the final analysis. The most common barriers were membership criteria (n = 11 of 23, 48%), such as being a student or staff member of a university; limited time availability (n = 8, 35%), such as being a recurring event but not currently offered; and paywalls (n = 7, 30%). The average paywall amount was $99 USD, the cheapest being $10 USD and most expensive being $185 USD (n = 5). A full PRISMA flow diagram with exclusion reasons is reported in Figure 1.

**Figure 1.**
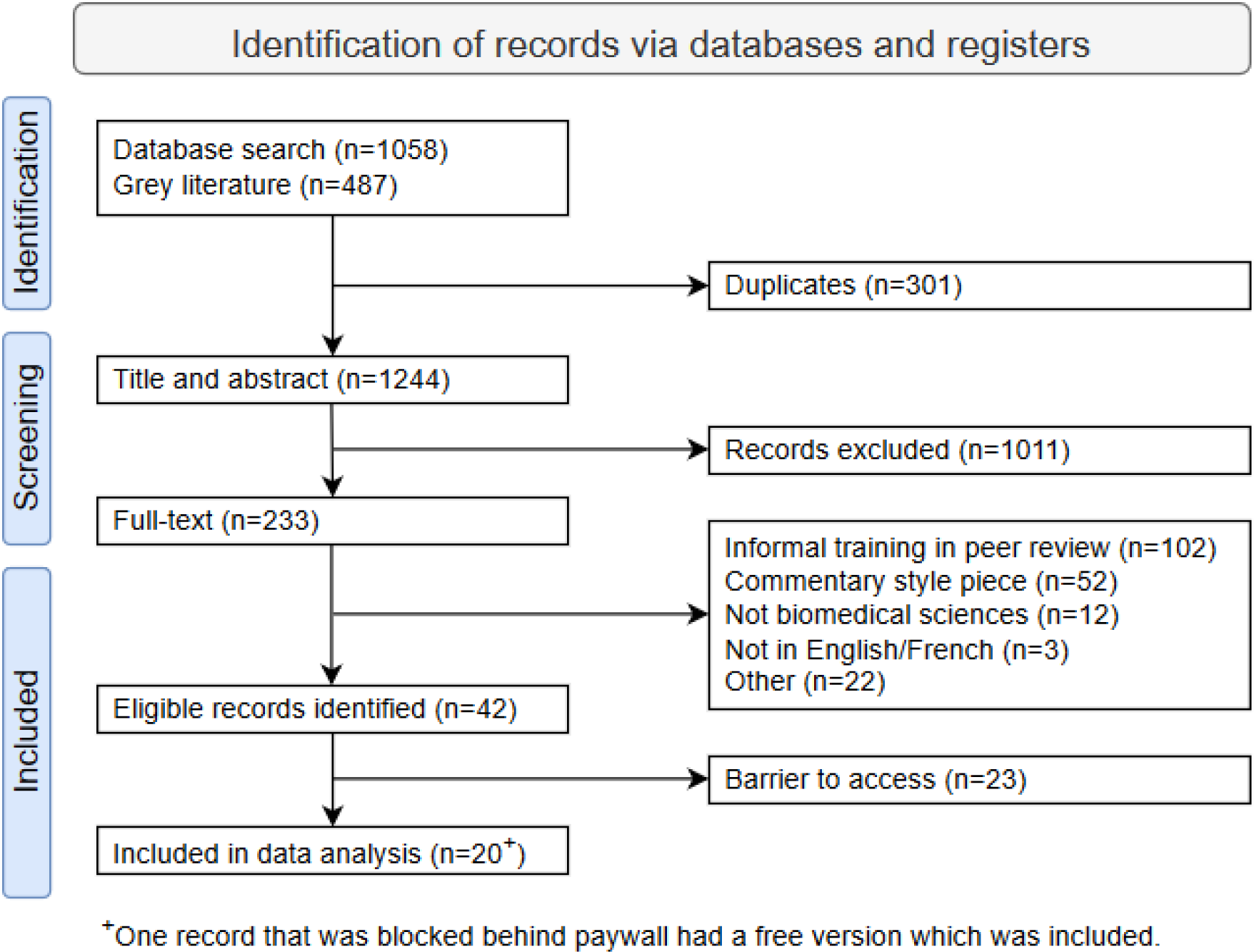
PRISMA flow diagram of included studies.

### 3. Training characteristics

In total, there were 20 sources that met the criteria of being fully openly available online training sources for peer review and were included in the data analysis. Full characteristics are listed in Table 1.

**Table 1.** Characteristics of training sources (oldest to most recent) Abbreviation: NR, not reported; ECR, early career researchers.

| Author, Year | Title / Link | Country | Organization | Target audience | Funding | Format | TTC | Proof of completion | Languages |
| --- | --- | --- | --- | --- | --- | --- | --- | --- | --- |
| Dickersin, 2012 | Free Online Course on Journal Peer Review | USA | Other | Other | Public | Online module | 4-8h | No | No |
| Chandran, 2013 | Peer Review of Manuscripts: An Online Training Module | USA | Academic | All researchers | NR | Online module | 0.5-1h | No | No |
| Sainani, 2017 | Doing a peer review | USA | Academic | All researchers | NR | Online video | 10-30 min | Other | Multiple (subtitles) |
| Veis, 2018 | Journal Peer Review: Tips for Writing an Effective Evaluation | USA | Journal | Journal reviewers | NR | Webinar | 0.5-1h | No | No |
| Foster, 2018 | Open Peer Review | Multiple | Other | No | NR | Online module | 0.5-1h | Other | No |
| Tokalić, 2018 | A peer review card exchange game | Croatia | Academic | All researchers | Private | Card game | 10-30 min | No | No |
| Chauvin, 2019 | COBPeer training module | France | Academic | ECR | None | Online module | 10-30 min | Other | No |
| Lovick, 2020 | How to Be A Peer Reviewer | USA | Publisher | No | NR | Webinar | 0.5-1h | No | No |
| Stiller-Reeve, 2021 | How to master peer review | Germany | Publisher | No | NR | Webinar | 1-2 hours | No | No |
| Elsevier, 2021 | Certified Peer Reviewer Course | Netherlands | Publisher | No | NR | Online module | 4-8 hours | Yes | Chinese |
| Marshall, 2021 | Peer Reviewer Training - How to Get Published | Multiple | Other | No | NR | Webinar | 1-2 hours | No | Chinese |
| Taylor & Francis, 2022 | Excellence in Peer Review: Expert Peer Review Training | Multiple | Publisher | All researchers | NR | Online module | 1-2 hours | Yes | No |
| ASHA Journals | The Peer Review Excellence Program | USA | Publisher | Journal reviewers | NR | Online module | 0.5-1h | No | No |
| The BMJ | Reviewer training materials | UK | Journal | Journal reviewers | NR | Website of resources | 0.5-1h | No | No |
| Nature Masterclass | Focus on Peer Review — free course | Germany | Publisher | ECR | NR | Online module | 2-4 hours | Yes | No |
| Optica | Reviewer certification | USA | Publisher | Journal reviewers | NR | Online module | 0.5-1h | Yes | No |
| Springer Nature | How to Peer Review | Germany | Publisher | All researchers | NR | Online module | 10-30 min | No | No |
| University of Manchester | My Research Essentials: Peer Review | UK | Academic | All researchers | NR | Online module | 10-30 min | No | No |
| Web of Science | An Introduction to Peer Review | UK | Other | ECR | NR | Online module | 10-30 min | Yes | No |
| Wolters Kluwer | Peer Reviewer Training Course | Netherlands | Publisher | All researchers | NR | Online module | 2-4 hours | No | No |

#### Author information, publication date and funding disclosures

Half of training sources did not have clear author information listed (n = 10, 50%). Similarly, most sources did not provide funding disclosures (n = 17, 85%). Eight sources (40%) did not report the publication date. Half of the sources (n = 10, 50%) were created in the last five years. Two (10%) were created prior to 2017.

Countries that created the most training sources include the United States (n = 7, 35%), United Kingdom (n = 3, 15%), and Germany (n = 3, 15%). Organizations that created training were most often publishers (n = 9, 45%) and academic institutions (n = 5, 25%).

#### Language and translation

All online sources were primarily published in English. Three (15%) had additional language options; one which was fully delivered in both English and Chinese (Mandarin), one which provided virtual workshops in either English or Chinese (Mandarin), and one which provided video subtitles in other languages.

#### Training format, target audience and approximate time for user to complete training

Most training sources were online modules (n = 13, 65%). Other formats included recorded webinars (n = 4, 20%), websites of resources (n = 1, 5%), an asynchronous online video (n = 1, 5%), and a game (n = 1, 5%).

Defined target audiences included all researchers (n = 6, 30%), reviewers at the journal which created the training (n = 5, 25%), and early career researchers (n = 3, 15%). Five sources (25%) did not define the target audience.

All training material took at least 10 minutes to complete. Most training material could be completed in less than one hour (n = 13, 65%), six (30%) could be completed in under half an hour and seven (35%) between half an hour and one hour. Three (15%) took 1-2 hours to complete, two (10%) took 2-4 hours and the two (10%) longest took 4-8 hours. Five sources (25%) were part of a larger course or curriculum that did not solely focus on peer review.

#### Proof of completion

Most training sources did not provide any proof of completion such as a certificate or badge (n = 12, 60%). Five sources (25%) provided certificate of completion. One source (5%) provided a certificate only after purchase of the full course. Other incentives for completion included CME credits (n = 1, 5%) and a badge (n = 1, 5%).

### 4. Content analysis of the training sources included

We assessed the 20 included sources for predefined content areas. A full list is included in Table 2 and depicted graphically in Figure 2.

**Table 2.**
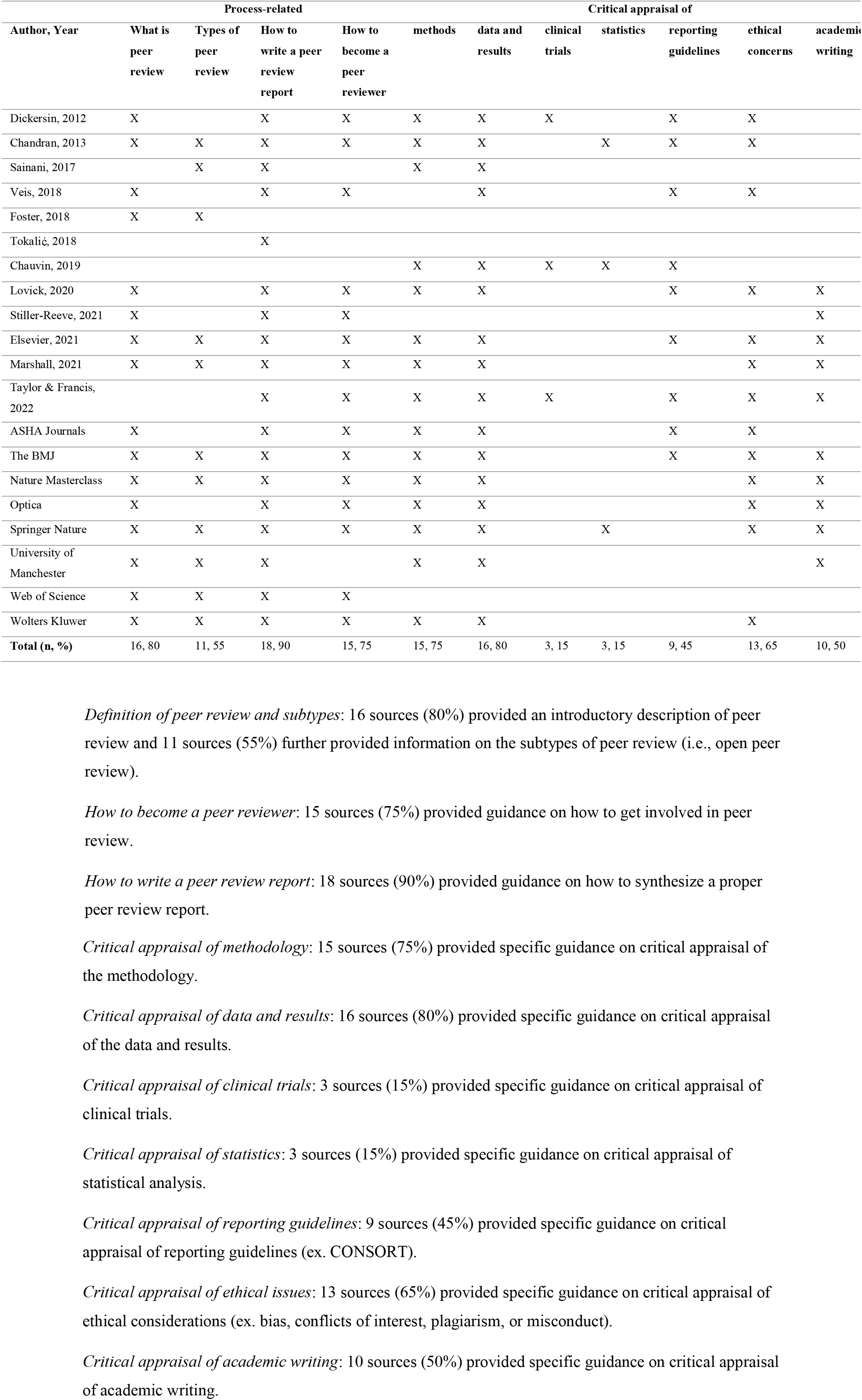
Content analysis of included topics.

**Figure 2.**
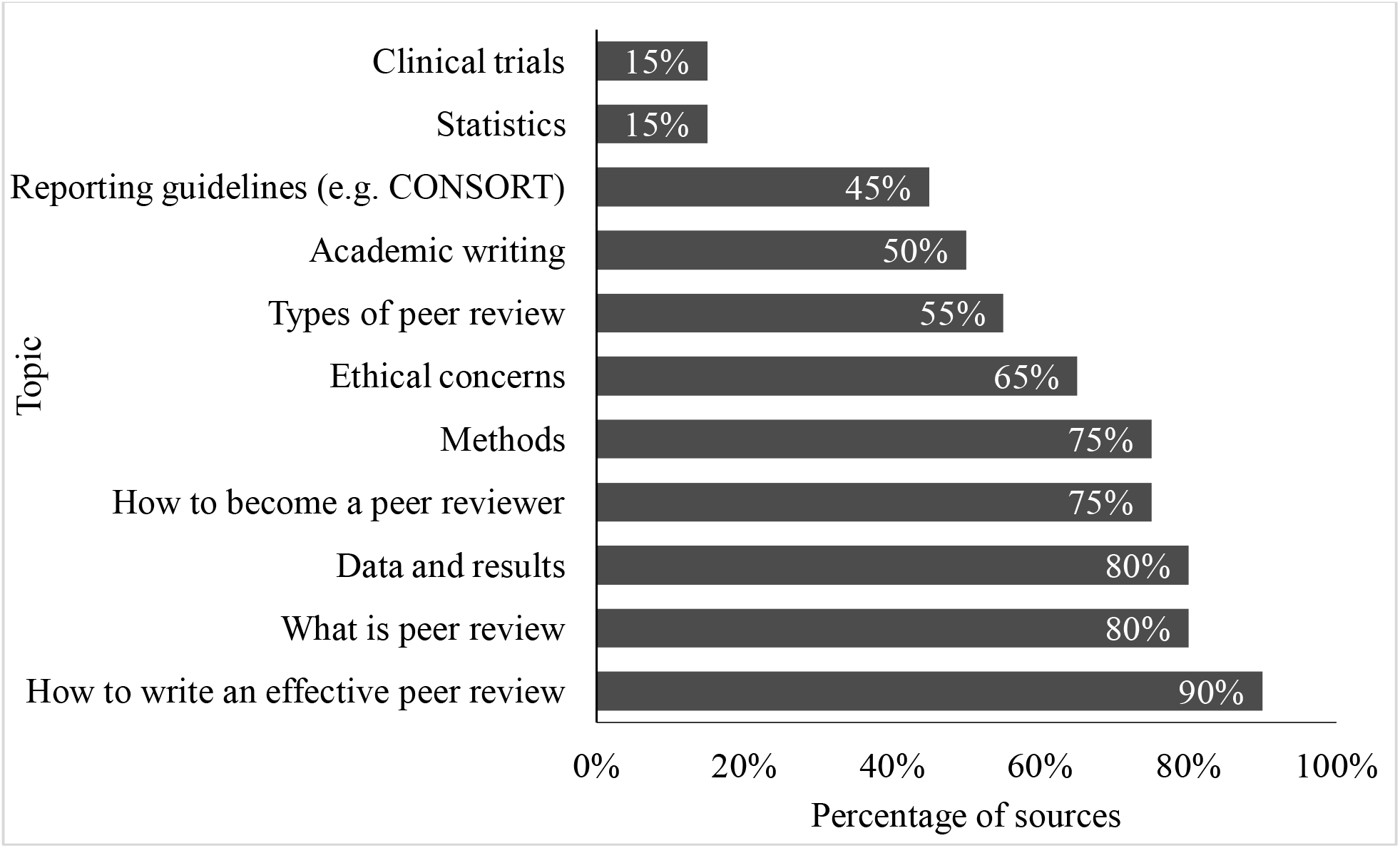
Content analysis of covered topics expressed as percentage of all included sources.

### 5. Risk of bias assessment

Overall, four training sources (20%) were assessed as evidence based. Full risk of bias items for each reference can be found in Table 3.

**Table 3.**
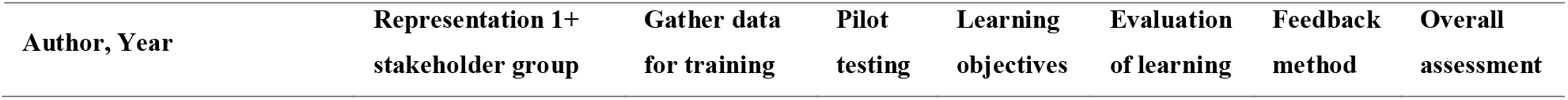
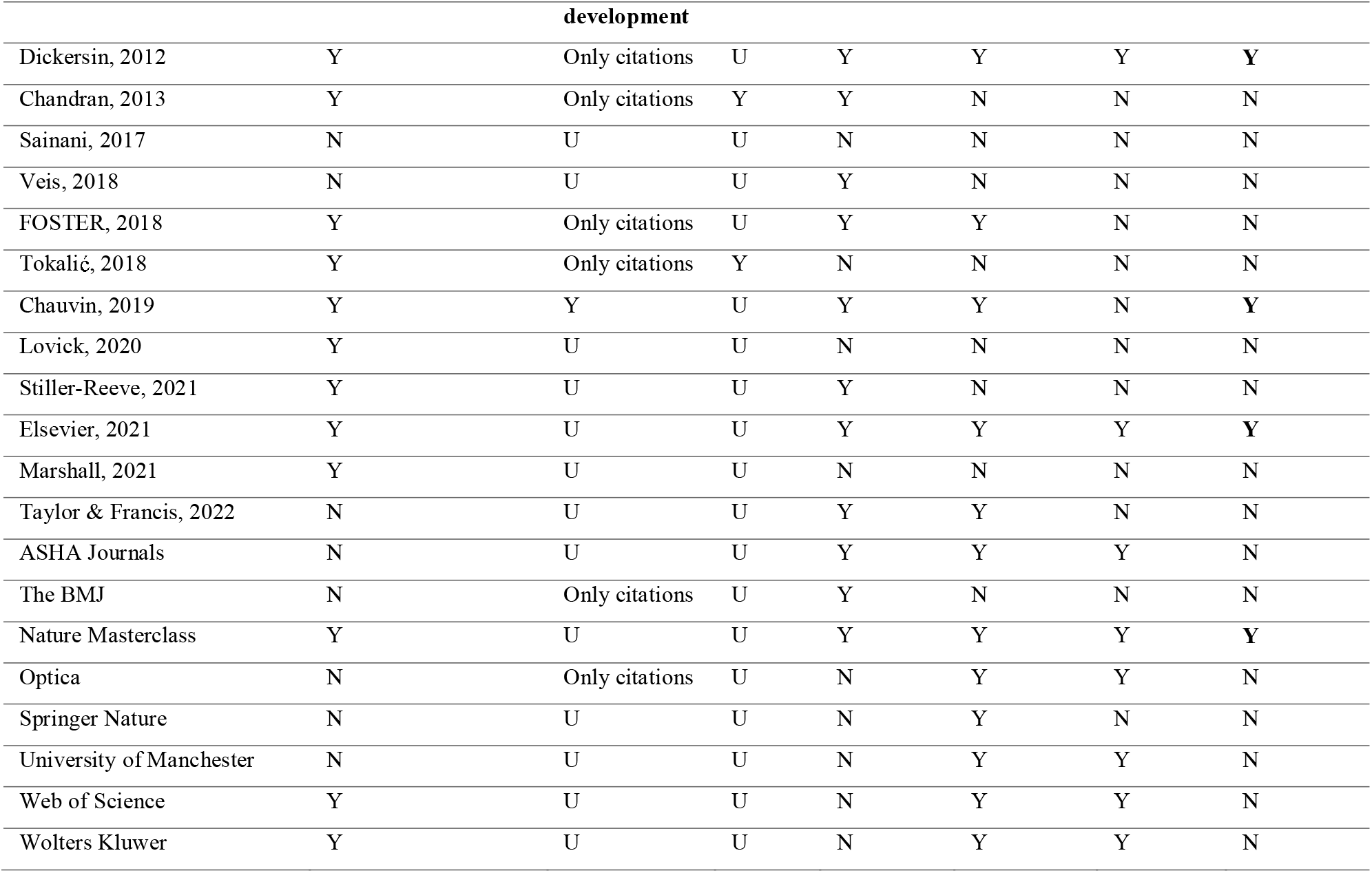
Internal validity tool of included sources.

Based on author affiliations, twelve training sources (60%) involved more than one stakeholder group in the creation or delivery (low risk of bias). Thirteen sources (65%) did not clearly report data gathering prior to the creation of the training material (unclear risk of bias). Eighteen sources (90%) did not clearly report pilot testing of the training (unclear risk of bias). Eleven training sources (55%) had learning objectives (low risk of bias). Twelve training sources (60%) provided a method for which a user could test their learning such as an online quiz or practice peer review sample (low risk of bias). Eight sources (40%) provided a method for users to provide feedback on the training material (low risk of bias).

## DISCUSSION

In our comprehensive search of the literature, we identified 43 training opportunities in manuscript peer review in the past 10 years, of which less than half were openly accessible online. Of the 20 included sources, more than half were online modules and could be completed in less than one hour. Overall, based on our assessment of internal validity, only four sources met our criteria of evidence-based.

For such a common activity, we were surprised by the small number of openly accessible online training sources in manuscript peer review. This is consistent with the findings of other previous studies^21^ as well as our own recently conducted survey of biomedical researchers in which most respondents indicated they had never received peer review training. Additionally, in our survey, being unable to find accessible training was identified as a barrier to pursuing training for most respondents^55^.

While the training sources that required less than an hour to complete may act as an appropriate introduction to peer review, it is not clear how much training can be acquired in such a short period of time. The three training sources that had the longest time to completion were additionally evaluated as evidence-based using our internal validity tool. Furthermore, this could explain why more complex topics, such as clinical trials, statistics and reporting guidelines were less commonly covered. In our recently conducted survey, the most desired training format was a full online course of greater than six sessions. Additionally, more complex topics, such as statistical analysis, were highly desired. We believe something more fulsome in content and time may result in higher quality peer reviewers and address existing knowledge gaps.

A lack of training in peer review may help to explain why there continues to be an abundance of low-quality research published^56^. Despite spending upwards of 100 million hours annually completing peer review^57^, it is unfortunate that more research has not examined ways to codify training and professionalism in peer review^19,21^. The scholarship around peer review appears to focus on ways to extend peer reviewing, such as to preprints^58^, or future innovations in peer review^1,7^. Did we get ahead of ourselves? How can we advance any aspect of peer review without getting the fundamentals right, namely, appropriate training?

Our systematic review is not without limitations. We selected the introduction of Publons training as our start date. It is possible that online training existed prior to this date. Nevertheless, the present results provide a comprehensive assessment of online training in the last decade. We only sought online openly accessible training resources, which is not the only training provided to peer reviewers. Be that as it may, in an era of equity, diversity, and inclusiveness, we think freely available opportunities provide the broadest opportunity for training, globally. Finally, it is unclear what risk of bias assessment is appropriate for methodological reviews, such as this one. We believed it best to use an assessment process used previously, rather than forego attempting to evaluate internal validity.

## CONCLUSION

Future collaborative efforts should focus on the creation of a more unified manuscript peer review training program. To meaningfully promote improving peer review skills, publishers could require all new peer reviewers to complete training and certification prior to peer reviewing. Furthermore, manuscript peer review is a global activity and training must be available to everyone interested in developing skills to complete high quality peer review. Special attention should be given to making training more accessible, equitable and inclusive.

## Data Availability

All data produced are available online on Open Science Framework.

https://osf.io/uac3k/

## Appendix 1: OVID search strategy

Database: Ovid MEDLINE(R) ALL <1946 to July 09, 2021>

1. ((peer review? or peer reviewer? or peer reviewing) adj2 (train* or curriculum* or education* or workshop* or module* or tutorial* or toolkit* or guide* or learn* or course* or webinar* or mentor* or lesson* or teach* or instruct*)).tw,kf. (356)
2. Curriculum/ or Computer-Assisted Instruction/ or Inservice Training/ or Education, Professional/ or Interactive Tutorial/ (109923)
3. Peer Review, Research/ (7055)
4. (peer review? or peer reviewer? or peer reviewing).ti. (4762)
5. (peer review? or peer reviewer? or peer reviewing).ab. /freq=2 (1968)
6. or/3-5 (11424)
7. 2 and 6 (117)
8. 1 or 7 (451)
9. limit 8 to yr=“2012-current” (201)

## Appendix 2: Grey literature search strategy

- Grey literature searches were done using the same inclusion and exclusion criteria as the OVID search.
- Forward and backward citation analysis of articles included at the data extraction stage was done using Web of Science on October 12, 2021.
- University library websites were searched on October 12, 2021. The Shanghai Ranking of World Universities 2020 (https://www.shanghairanking.com/rankings/arwu/2020) was used to identify the top 10 universities in each world region (Africa, Americas, Asian/Oceania, Europe). As our author group is based in Canada, we additionally searched the top 15 (U15) Canadian research university library sites on October 4, 2021.
- To better cover training material created by journals, we searched the top 6 biomedical publishers by journal count (Taylor & Francis, Elsevier, Springer, Wiley, SAGE, Wolters Kluwer Health), as well as publishers identified as having an interest in peer review (BMJ, AMA, eLife, PLOS, Frontiers) on October 12, 2021.
- Google (Oct 25, 2021) and YouTube (Oct 18, 2021) search of “peer review training” and “peer review course” limited to post-2012. Searches were done in an incognito browser. The results of each search were recorded, excluding search results from a publisher and replacing them until the first 100 results were recorded.
- Preprint server search using “peer reviewer training” on: OSF Preprints, bioRxiv, MediArXiv, EdArXiv, PeerJ, Preprints.org, PsyArXiv on October 12, 2021.
- Peer review groups/events that had a website were also be searched on October 12, 2021: EQUATOR Network (equator-network.org/), Publons (publons.com), PEERE (peere.org), and Peer Review Congress (peerreviewcongress.org/).

## Appendix 3: Screening form

Level 1 (title/abstract):

1. Does this document describe training for peer reviewers of scientific documents? (yes/unclear; no)

If yes/unclear, move to Level 2 (full text):

1. Does this document describe training for peer reviewers of scientific documents? (yes/unclear; no)
2. Is there any reason to exclude the document? (yes/no)
  a. If yes, please specify:
    i. Not in English/French
    ii. Not published on or after January 1^st^, 2012
    iii. Not openly available
    iv. Other, please specify:___

## Appendix 4: Data extraction form

| Item # | Item description | Answer options [*in italics, further instruction for extractor*] |
| --- | --- | --- |
| 1 | DOI | *If no DOI, use website URL* |
| 2 | Author | *Extract the first listed corresponding author. If not, corresponding author is indicated, extract the first listed author. If reported as a group, extract group name. If not reported, write ‘not reported’.* |
| 3 | Year of publication | *Use the most recent year stated on the document* |
| 4 | Country where training was developed | *If multiple countries are listed, specify 'multiple countries'* |
| 5 | Organization that developed the training | *If unclear, extract based on author from 2.*<br>*If multiple organizations are listed, specify 'multiple organizations'* |
| 6 | Target audience | *i.e., was the training created for peer reviewers of a specific journal/publisher, university?*<br>Yes, please specify: _____<br>No |
| 7 | Funding | Public<br>Private<br>Both private and public<br>None<br>Not reported |
| 8 | Training format | *Check all that apply.*<br>Online module<br>Website of resources<br>In-person workshop<br>Webinar/Zoom<br>Online video (asynchronous)<br>Other, describe: ____<br>Unsure |
| 9 | Is the training provided as part of a larger course/curriculum? | Yes, please specify: _____<br>No<br>Unsure |
| 10 | Does the training report the length of | Yes, please specify: ____<br>If not reported, make an estimate: ____ |
|  | time it takes? |  |
| 11 | Does the training provide proof of completion (i.e., certificate)? | Yes, please specify: ____<br>No<br>Not reported |
| 12 | Are there any barriers to access the training online? | *Check all that apply.*<br>No barriers<br>Paywall, please specify amount: ____<br>Registration<br>Membership, please specify for what: ____<br>Other, please specify: ____ |
| 13 | Is the training offered in any other languages? | Yes, please specify: ____<br>No<br>Unsure |
| 14 | What are the topics covered by the training? | *i.e., look at the sub-headings, table of contents if helpful*<br>*Check all that apply.*<br>• What is peer review<br>• Types of peer review (e.g., open peer review)<br>• How to write an effective peer review report<br>• How to become a peer reviewer<br>• Critical appraisal of...<br>○ Methodology (randomization, interventions, etc.)<br>○ Data and results<br>○ Clinical trials<br>○ Statistics<br>○ Reporting guidelines (ex. CONSORT)<br>○ Bias, conflict of interest, plagiarism, and misconduct<br>○ Academic writing<br>• Other, please specify: ____ |
| 15 | Does the document report to have evaluated the peer review training described? | Yes<br>No<br>If yes, what study design was used? (Observational study, randomized trial, experiment, other, please specify: ____)<br>If trial, briefly describe the participants and outcomes. |

## Appendix 5. Risk of bias form

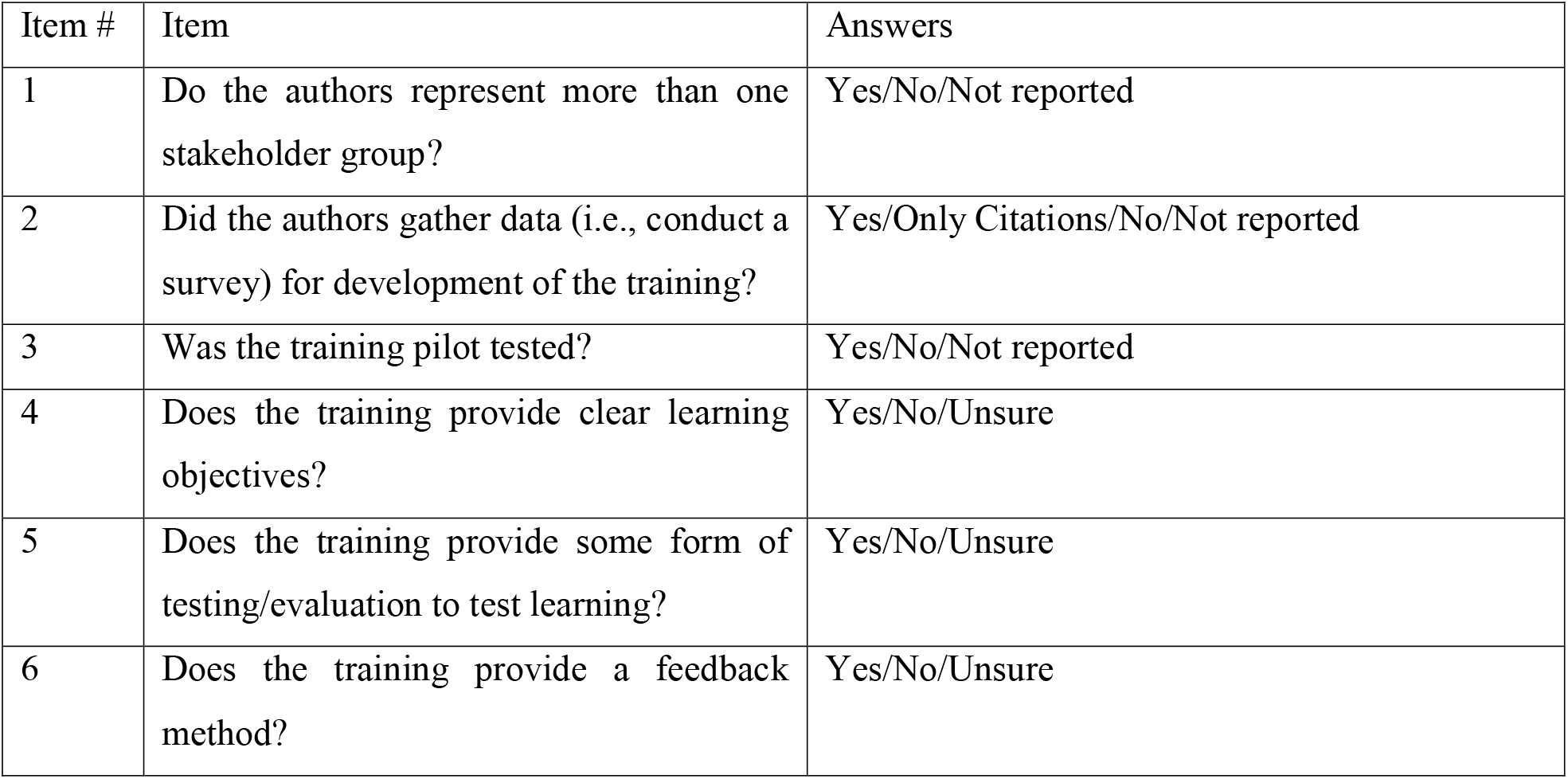

| Item # | Item | Answers |
| --- | --- | --- |
| 1 | Do the authors represent more than one stakeholder group? | Yes/No/Not reported |
| 2 | Did the authors gather data (i.e., conduct a survey) for development of the training? | Yes/Only Citations/No/Not reported |
| 3 | Was the training pilot tested? | Yes/No/Not reported |
| 4 | Does the training provide clear learning objectives? | Yes/No/Unsure |
| 5 | Does the training provide some form of testing/evaluation to test learning? | Yes/No/Unsure |
| 6 | Does the training provide a feedback method? | Yes/No/Unsure |

